# No difference in retinal fluorescence after oral curcumin intake in amyloid proven AD cases compared to controls

**DOI:** 10.1101/2021.10.07.21264420

**Authors:** Jurre den Haan, Frederique J. Hart de Ruyter, Benjamin Lochocki, Maurice A.G.M. Kroon, E. Marleen Kemper, Charlotte E. Teunissen, Bart van Berckel, Philip Scheltens, Jeroen J. Hoozemans, Aleid van de Kreeke, Frank D. Verbraak, Johannes F. de Boer, Femke H. Bouwman

## Abstract

**INTRODUCTION:** Previous work showed the *in-vivo* presence of retinal amyloid in AD patients using curcumin. We aimed to replicate these findings in an amyloid biomarker confirmed cohort.

**METHODS:** Twenty-six AD patients (age 66 (+9), MMSE≥17) and 14 controls (age 71(+12)) used one of three curcumin formulations: Longvida^®^, Theracurmin^®^ and Novasol^®^.

Plasma levels were determined and pre- and post-curcumin retinal fluorescence scans were visually assessed in all cases and quantitatively assessed in a subset.

**RESULTS:** Visual assessment showed no difference between AD patients and controls for pre- and post-curcumin images. This was confirmed by quantitative analyses on a subset. Mean conjugated plasma curcumin levels were 198.7 nM (Longvida^®^), 576.6 nM (Theracurmin^®^) and 1605.8 nM (Novasol^®^).

**DISCUSSION:** We found no difference in retinal fluorescence betweenamyloid confirmed AD cases and control participants, using Longvida^®^ and two additional curcumin formulations. Additional replication studies in amyloid confirmed cohorts are needed to assess the diagnostic value of retinal fluorescence as an AD biomarker.

## Background

Alzheimer’s disease (AD) research over the last decades enabled clinicians to diagnose AD not only in the dementia end-stage but also in the prodromal and preclinical stages using biomarkers for amyloid, tau and neurodegeneration [1, 2]. AD pathology starts 15-20 years before symptom onset and is reflected by change of biomarkers in the preclinical stage [1]. This time-interval provides a window of opportunity for disease modifying drugs to potentially halt disease progression towards the end stage of AD: dementia [3]. As currently used biomarkers are time consuming, expensive and/or invasive, non-invasive easily accessible biomarkers are urgently needed to diagnose AD in the earliest stages, enabling timely selection of patients for trials and future medication.

The retina is an extension of the central nervous system and might provide such a biomarker. As such the retina is increasingly studied for manifestations of AD as potential non-invasive AD biomarkers. For example, retinal thinning, changes in retinal vasculature and peripheral drusen have been described in AD patients [4-6]. More specifically related to AD pathology, three small studies reported visualization of retinal amyloid beta deposits *in-vivo* using curcumin, a polipotent polyphenol with fluorescent properties that binds to amyloid in post-mortem brain tissue [7-9]. The presence of retinal amyloid beta deposits is however not unequivocally proven, since we and other groups could not confirm amyloid beta deposits in post mortem retinal tissue [10-14]. Moreover, curcumin is known for its low bioavailability, and while several commercially available formulations enhancing curcumin plasma levels are on the market, so far only one formulation (Longvida^®^) has been reported to visualize retinal amyloid[7].

In the current study, we used three different curcumin formulations as labeling fluorophores in-vivo. Using a targeted fluorescence approach, we aimed to visualize retinal amyloid in a well-characterized and amyloid biomarker confirmed AD cohort.

## Methods

### Participants

We enrolled 26 patients with AD (MMSE score ≥17) and 14 controls (MMSE ≥27) from two different cohorts – the Alzheimer Dementia Cohort (ADC) and the EMIF-AD PreclinAD study. All subjects underwent extensive screening according to a standardized protocol described elsewhere [15, 16]. All patients fulfilled NIA-AA criteria of AD with amyloid biomarker confirmation through either CSF amyloid beta_1-42_ (Aβ_42_) analysis or amyloid-PET [1]. ADC controls had subjective cognitive decline, defined as cognitive complaints without objective cognitive impairment on neuropsychological exam, no signs of neurodegeneration on neuroimaging, and absence of AD pathology based on CSF biomarkers and/or amyloid-PET. Exclusion criteria for all participants were ophthalmological conditions interfering with retinal scan quality, e.g. diabetic retinopathy, glaucoma or moderate/intermediate age related macular degeneration.. In addition, we excluded subjects with ischemic stroke and/or mild to severe white matter hyperintensities on MRI, operationalized as a Fazekas score >2. We excluded two AD patients; one because of problems with eye fixation due to visuoperceptive dysfunction and one because of participation in a drug trial with disease modifying drugs. We excluded two controls; one due to problems finishing the scan protocol because of dry eyes and one was found to have glaucoma. Three AD patients were lost to follow up. This study was designed and conducted according to the Declaration of Helsinki and the study protocol was approved by the Ethical Committee of the VU University Medical Center. All patients gave their written informed consent in the presence of their caregiver.

### Amyloid biomarker assessment

Between 2016 and 2018, CSF concentrations of Aβ_42_, total tau, and tau phosphorylated at threonine 181 (pTau_181_) were measured using Innotest ELISAs (Fuijirebio, Ghent, Belgium). Between 2018 and 2020, these CSF biomarkers were assessed using Elecsys Aβ_1–42_ CSF, Elecsys total-tau CSF, and Elecsys pTau_181_ CSF electrochemiluminescence immunoassays (Roche Diagnostics, Basel, Switzerland). For comparability Innotest CSF values were converted using previously published conversion formulas [17]. A pTau/ Aβ42-ratio≥0.020 was considered an AD profile [18].

Amyloid-PET scanning was performed with either ^18^F-Florbetaben (NeuraCeq), ^18^F-Florbetapir (Amyvid) or 18F-Flutemetamol (Vizamyl) tracers [19-21]. An experienced nuclear physician (BvB) who completed training for all radiotracers visually assessed images of amyloid-PET scans as positive or negative.

### Ophthalmological assessment

Subjects underwent the following general eye examination: best corrected visual acuity (VA), intraocular pressure (IOP) using non-contact tonometry, and slit-lamp examination of the anterior and posterior segment, followed by administration of tropicamide 0.5% to dilate the pupil for optimal retinal imaging. An experienced ophthalmologist (FDV) interpreted all examinations.

### Heidelberg HRA SLO imaging

Retinal fluorescence imaging was performed with a Heidelberg Engineering Spectralis Spectral Domain Scanning Laser Ophthalmoscope, at a 486nm wavelength (blue autofluorescence) laser source to excite fluorescence, in combination with a long-pass 500 filter (high transmission in 498-760nm and 800-835nm). Optical resolution was 12 µm with a 55° lens, and 6 µm with a 30° lens. For each final image an average of 50 frames was used with a sensitivity >90.

### Curcumin administration and timing of fluorescence imaging

Three different oral formulations of curcumin were used:

1. Longvida^®^ Solid-Lipid Curcumin Particle (Verdure Sciences^®^), was given in a dosage of 4000 mg per day for 10 consecutive days to 14 AD patients and 12 controls [7]. Here we used a 30° lens to acquire images in six regions of interest (ROI) (central macula, superior, temporal, inferior, superior-temporal and inferior-temporal) at baseline and 2-4 hours after the 10th day of curcumin intake (Figure 1A-B).
2. Theracurmin^®^ (Theracurmin; Theravalues, Tokyo, Japan), was given in a dosage of 180 mg for 5 consecutive days to 7 AD patients and 2 controls [22, 23]. Here we acquired images using a 30° lens in three ROI’s (central macula, optic nerve head (ONH) and temporal) at four different time points: i) at baseline, ii) after four days of curcumin intake, iii) 1 hour after the fifth dose, and iv) 2 hours after the fifth dose (Figure 1 A-B).
3. Novasol^®^ (AQUANOVA AG, Darmstadt, Germany), was given in a dosage of 300 mg for 4 consecutive days and a final dose of 500 mg on day 5 to 5 AD patients [24, 25]. With Novasol we acquired images using a 55° lens in six ROI’s (central macula, superior, temporal, inferior, superior-temporal and inferior-temporal) at baseline and 2 hours after the fifth dose (Figure 1A-B).

**Figure 1.**
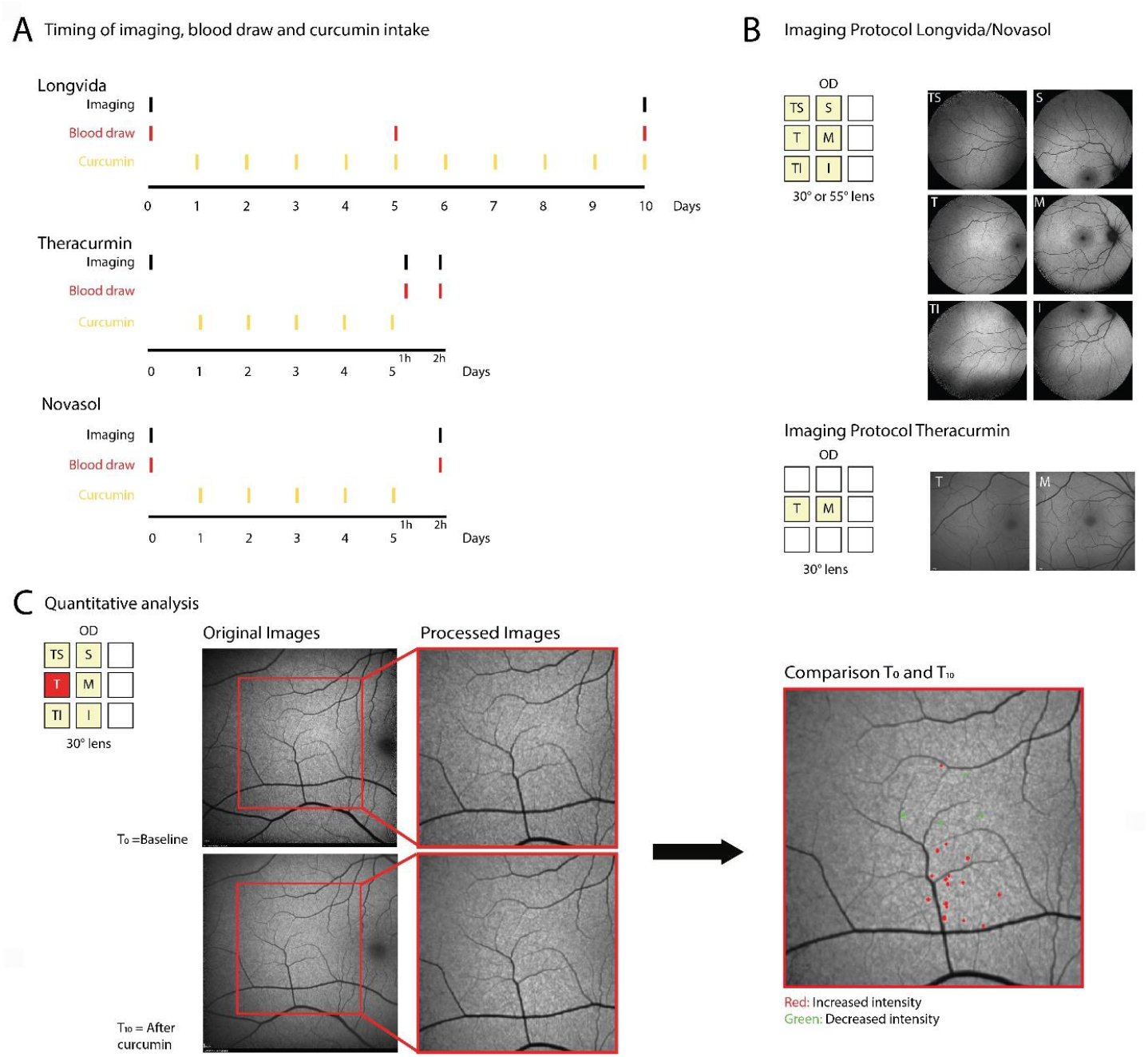
Visual representation of study protocol. A. Timing of imaging, blood draw and curcumin intake of cohort 1 (Longvida^®^, 4000 mg for 10 days), cohort 2 (Theracurmin^®^, 180 mg for 5 days) and cohort 3 (Novasol^®^, 300 mg for 4 days, 500 mg for 1 day). B. Imaging protocol of cohort 1, 2 and 3. C. Quantitative analysis.

### Retinal fluorescence image analysis

Baseline and post-curcumin-administration images were assessed both visually and quantitatively. Visual assessment was performed by an experienced ophthalmologist, masked for diagnosis and curcumin treatment (FDV). Baseline and post-curcumin-administration images of AD patients were compared for the Novasol cohort and also between AD and control subjects for the Theracurmin and Longvida cohort. To support the visual assessment, we used the Longvida^®^ cohort for quantitative analysis, allowing comparison to previous publications using Longvida^®^ as curcumin derivative. Images of the right eye with sufficient image quality (scans that were out of focus, had extensive shading or extreme low/high contrast hampering co-registration were excluded) were assessed for quantitative analysis (Figure 1C). Retinal fluorescence imaging yielded 8-bit grayscale images sized 1536 × 1566 or 768 × 798 pixels with a resolution of 6 or 12 µm respectively per pixel. The non-normalized, uncompressed images were compared between time points using MATLAB 2020b (MathWorks). An image pair of two time points of similar ROI’s were cropped to a size of 1430 × 1430 or 715 × 715 pixels to remove the manufacturer logo and blurry edges. In the case any image pair contained high resolution images (1430 × 1430) the images were down-sampled to match the size of 715 × 715 pixels. Afterwards, both images were flat-field corrected to compensate for any illumination and shading effect. A registration algorithm (feature-based registration (SURF: Speeded-Up Robust Features) with affine transformation, allowing for rotation) was used to register matched images [26]. If an automatic match was not obtained, the software interrupted and allowed for manual picking of identical points of interest in both images and creating a registration match based on the user’s input (using the same registration algorithm). Registered images were cropped to a size of 512 × 512 pixels, ensuring that both images overlapped within the imaged field of view. Brightness histograms of both images were obtained and matched (using a cubic polynomial) to the histogram of the baseline image, resulting in images with similar contrast and overall brightness. We defined focal retinal hyperfluorescence, by selecting areas of ≥4 adjacent pixels that represented the 10% highest pixel value in the post-curcumin image. These hyperfluorescence spots were labeled, and their area and brightness measured. The same properties were measured in the baseline image, using the exact same spots (as identified in the post-curcumin image). Afterwards, the brightness difference between each hyperfluorescence spot was calculated.

### Curcuminoids plasma level analysis

Blood was drawn in heparinized tubes that were directly kept in aluminum foil on ice, to avoid degradation by light and temperature. Within 30 minutes, plasma and serum were separated and plasma was stored at minus 80°C in the Amsterdam UMC Biobank. High Performance Liquid Chromatography-Tandem Mass Spectroscopy (HPLC-MS-MS) analysis was performed to measure different curcuminoids: curcumin, demethoxycurcumin, bisdemethoxycurcumin and tetrahydrocurcumin. Each sample was treated, in duplicates, with and without β-glucuronidase after which unconjugated and conjugated curcuminoids were quantified. Treatment with β-glucuronidase hydrolyzes the conjugated curcuminoids leading to an increase in unconjugated curcuminoids, whereas untreated samples were quantified on unconjugated curcumin at that point of time. Our validated method of analysis can be described, in short, as follows: Liquid-liquid extraction with Tert Butyl Methyl Ether (TBME) was used to extract the compounds from the plasma [27]. The HPLC-MS/MS system consisted of an Ultimate 3000 autosampler and pump, both of Dionex, connected to a degasser from LC Packings. The autosampler, with a 100 µL sample loop, was coupled to an Sciex API4000 mass spectrometer. A total of 50 µL sample was injected after which separation of the analytes was performed with an Agilent column 2.1×100mm packed with material of Zorbax Extend 3.5 µm C-18. The flowrate was 0.200µL/min and the dual gradient mobile phase consisted of A: ultra-purified H2O with 0.1% formic acid and B: MeOH 100%. The applied gradient profile started at 50:50 A:B and increased linear to 95% B in 3.0 minutes. During 6 minutes a 5:95 A:B level is continued, after which it returned in 0.2 minutes to 50:50. Afterwards the system was equilibrated during 6 minutes at the starting level. Throughout the liquid-liquid extraction and HPLC-MS/MS the potential influence of light brought back to a minimum by working in a dark environment. Each analytical run included a set of freshly prepared calibration samples containing all compounds in the validated range of 2 nM to 400 nM. Each compound was validated with its own deuterated internal standard.

### Statistical analysis

SPSS version 26.0 (IBM, Armonk, NY, USA) was used to assess group differences in demographics, curcumin data and quantitative retinal fluorescence. Chi squared test was used for dichotomous variables, independent sample T-test was used for continuous variables that were normally distributed and Mann Whitney U Test was used for non-normal distributed variables. Quantitative comparison of focal retinal hyperfluorescence between AD patients and controls was assessed by calculating the number of focal retinal hyperfluorescent pixels defined as a value in the top 10% value range at baseline and after curcumin, for all retinal regions of the right eye. Between group differences were assessed using a Mann Whitney U test. Significance level for all tests was set at 0.05.

## Results

Group demographics of AD and control cases are shown in table 1, and supplemental table 1 shows data for each individual participant. The control group comprised more female participants. No differences were found for age between AD and control group. As expected, and by design, we found significant group differences for MMSE as well as CSF and amyloid-PET biomarkers. Table 2A shows the demographics of the sub group used for the quantitative analysis. No statistical group differences in age and sex were found.

**Table 1.** Demographics.

|  | **Alzheimer's**<br>**disease**<br>*n* = 26 | **Controls**<br>*n* = 14 | p-value |
| --- | --- | --- | --- |
| **Sex (F/M)** | 10/16 | 10/4 | .047<sup>a</sup> |
| **Age, mean (SD)** | 67 (9) | 71 (12) | .177<sup>b</sup> |
| **MMSE, median (IQR)** | 24 (21-26) | 29 (29-30) | .000<sup>c</sup> |
| **CSF Aβ, median (IQR)** | 541 (466-744) | 1328 (982-1551) | .002<sup>c</sup> |
| **CSF Tau, median (IQR)** | 320 (242-472) | 208 (123-322) | .060<sup>c</sup> |
| **CSF pTau, median (IQR)** | 34 (23-44) | 19 (9-30) | .035<sup>c</sup> |
| **CSF biomarker profile**<br>**(positive/negative)** | 20/0 | 3*/2 | .033<sup>a</sup> |
| **Amyloid-PET (visual read:**<br>**positive/negative)** | 17/0 | 0/13 | .000<sup>a</sup> |

CSF data (presented as pg/ml) was available in 20 AD cases and 5 controls.
Amyloid-PET data (visual read) was available in 17 AD cases and 13 controls. <sup>a</sup>Chi
squared test, <sup>b</sup>Independent sample T-test, <sup>c</sup>Mann Whitney-U test. \*3 controls had a
borderline positive biomarker profile, with a negative amyloid-PET. Abbreviations: SD
= standard deviation; MMSE = mini mental state exam; IQR = interquartile range;
pTau = phosphorylated tau; CSF = cerebral spinal fluid.

**Table 2.**
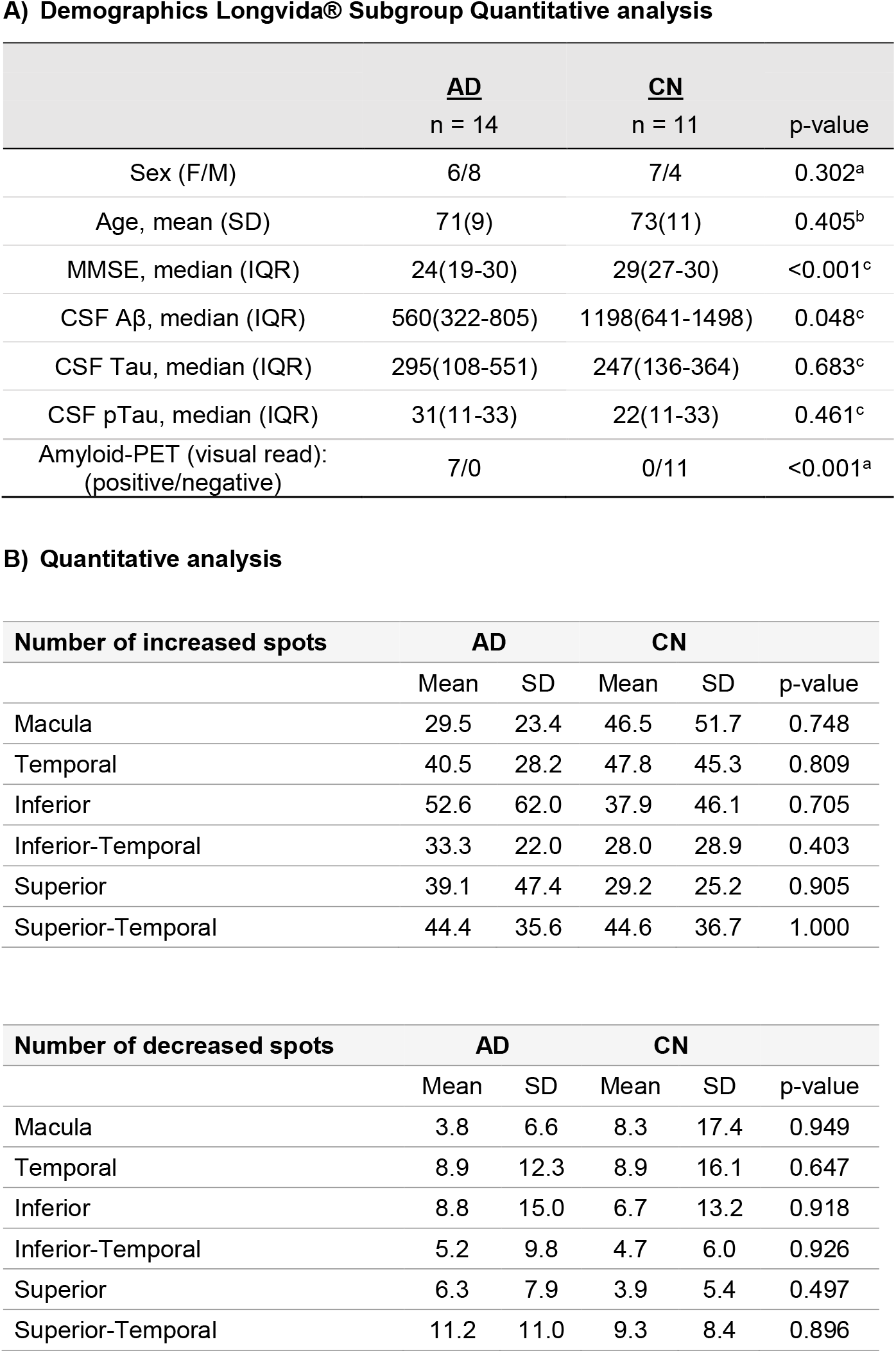
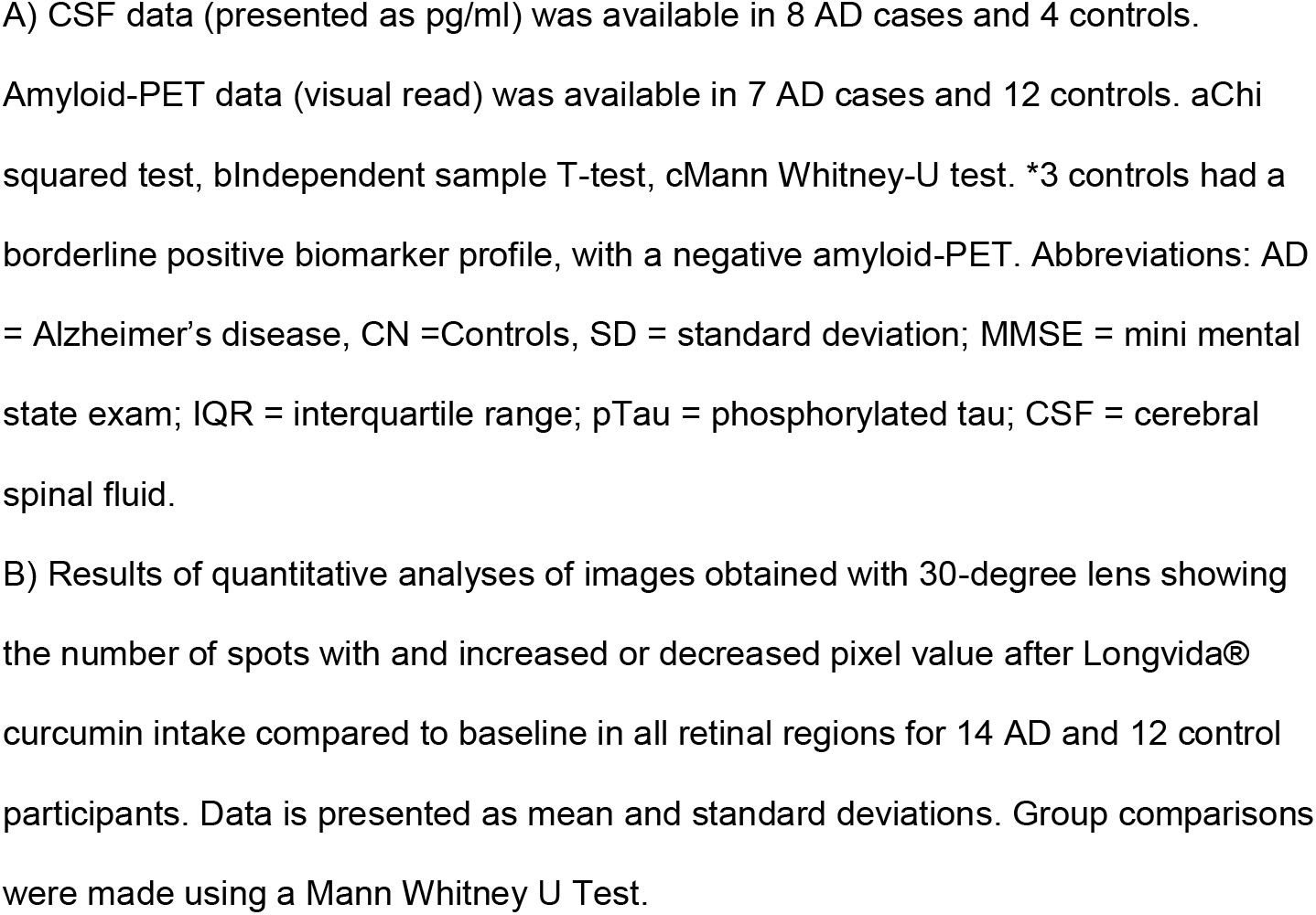

### Qualitative and quantitative retinal fluorescence analysis

No scans were excluded for the qualitative analysis. The qualitative assessment of images pre- and post-curcumin intake by an experienced ophthalmologist (FDV) masked for the clinical diagnosis did not show differences in focal hyperfluorescence in the ROI’s, for either AD patients (Longvida^®^, Theracurmin^®^ and Novasol^®^) or controls (Longvida^®^ and Theracurmin^®^)(Figure 2 and supplemental Figures 1 and 2). The Longvida^®^ cohort was used for quantitative analysis. For the quantitative analysis one out of 12 control cases (case #10, see supplemental Table 1) was excluded, as our algorithms were unable to find a good registration between time points. This applied to a total of 24 regions of a total of 10 patients, while the other regions (n=132) could be included in the analysis. All 14 AD cases that used Longvida^®^ were included in the quantitative analysis. While we did find an overall increased fluorescence after curcumin intake (data not shown), no differences were found between diagnostic groups in the number of spots with increased or decreased fluorescence after curcumin intake for all ROIs (all p-values>0.3) (Table 2B).

**Figure 2.**
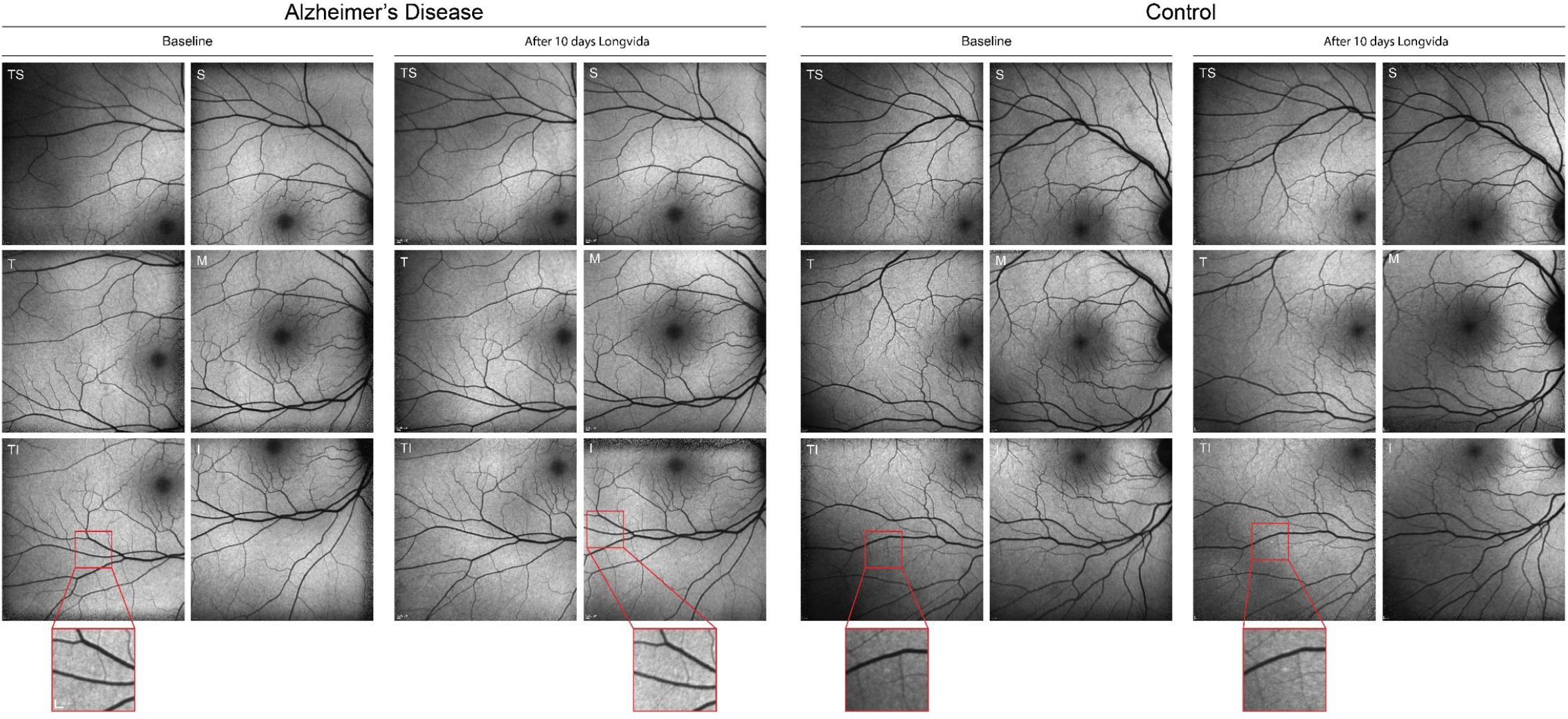
Pre- and post-curcumin retinal fluorescence images for AD and control participants using Longvida^®^. Pre-and post-curcumin retinal fluorescence images using blue auto fluorescence (λ=486nm) in 6 retinal regions in a representative AD patient and control. Magnifications show incidental focal hyperfluorescence, both on baseline and after curcumin in AD and control participants. Abbreviations: TS= temporal-superior, S= superior, T= temporal, M= macula, TI= temporal-inferior, I= inferior.

### Curcuminoids plasma level analysis

HPLC-MS/MS analysis of plasma samples showed detectable levels of free curcumin in blood around the detection level (2 nM) after Longvida^®^, Theracurmin^®^ or Novasol^®^ intake in all 40 subjects, however there were no significant differences between groups. After treating the samples with β-glucuronidase, catalyzing the separation of curcumin from glucuronide and sulfate conjugates, curcumin levels were higher (Table 3). The total mean of curcuminoids was 156.2 nM (±169.9) after Longvida^®^, 576.6 nM (±211.1) after Theracurmin^®^ and 1605.8 nM (±524.6 nM) after Novasol^®^ (Figure 3). There were no differences in plasma levels between diagnostic groups (p >0.3, data not shown).

**Table 3.**
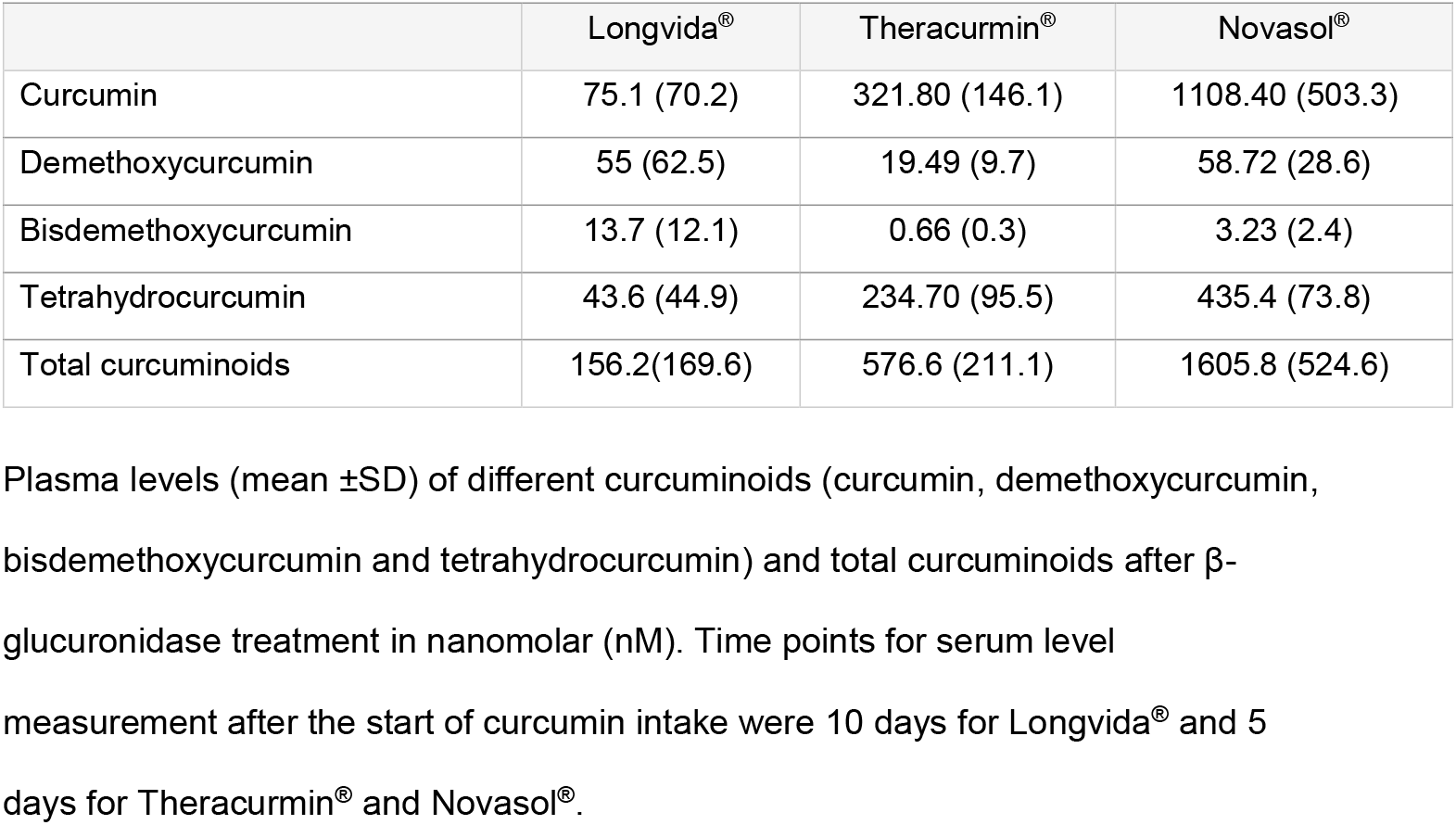
Curcuminoids plasma levels.

**Figure 3.**
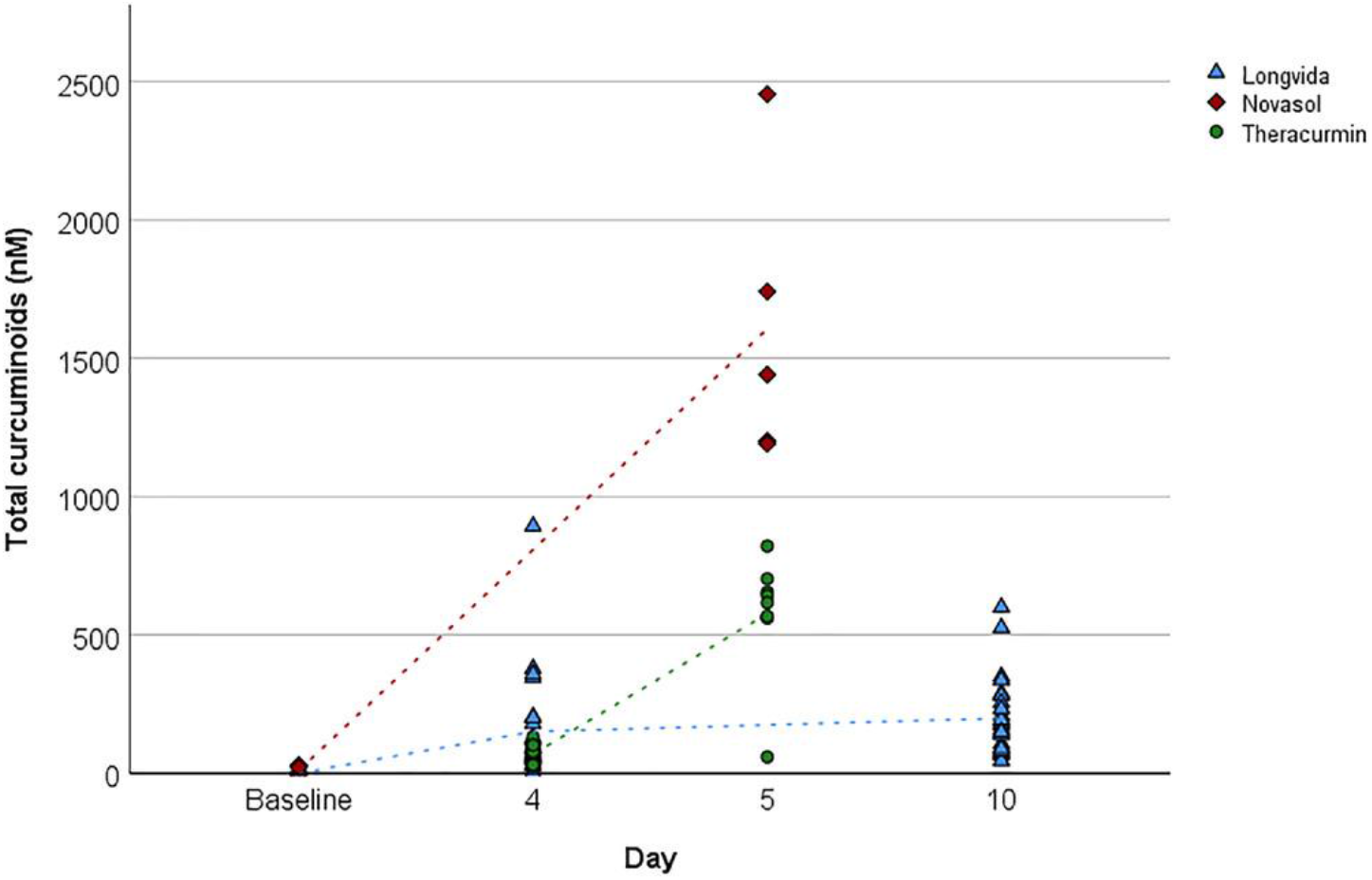
Plasma levels of total curcuminoids after Longvida^®^, Theracurmin^®^ and Novasol^®^ intake measured with HPLC-MS/MS. Overview of plasma levels of total curcuminoids after Longvida^®^, Theracurmin^®^ and Novasol^®^ intake. Curcumin, demethoxycurcumin, bisdemethoxycurcumin and tetrahydrocurcumin as measured with HPLC-MS/MS after β-glucuronidase treatment, summed as total curcuminoids in nanomolar (nM).

## Discussion

In this study we aimed to discriminate AD patients from controls with a targeted fluorescence approach of the retina using curcumin as fluorophore in a well-characterized AD biomarker confirmed cohort. Using three different curcumin formulations, we did not find differences in focal retinal hyperfluorescence before or after curcumin intake between diagnostic groups, despite an observed overall increase in retinal fluorescence after curcumin intake.

Our negative findings may be explained by a low intensity of the fluorescent signal resulting from (amyloid-bound) curcumin, insufficient binding of unconjugated curcumin to retinal amyloid, absence of retinal amyloid (in this subset of patients) or methodological limitations of our scanning and analysis methods.

Fluorescence of curcumin largely overlaps with autofluorescence of the retina. This might therefore mask subtle signals of curcumin fluorescence hypothesized to be the result of retinal amyloid (plaque) pathology, reported to be approximately 5-20µm in size [7]. At present no data is available describing the magnitude of fluorescent signal that can be expected from retinal amyloid relative to retinal autofluorescence. On visual inspection we could identify a certain number of hyperfluorescent spots (Figure 3, zoom-ins), however there was no difference between AD patients and controls in these spots, either in size or intensity, neither at baseline, nor after curcumin intake. To increase sensitivity for weak retinal fluorescent signal possibly overlooked by visual inspection, we performed a quantitative analysis on a subset of participants (14 AD, 11 controls) who received Longvida confirming findings of the visual inspection.

The absence of a fluorescent signal after curcumin intake could also be explained by insufficient curcuminbinding to retinal amyloid due to low plasma levels of unconjugated curcumin. After deconjugation using β-glucuronidase we observed high blood levels of total curcumin, in line with previous studies [22, 24]. In contrast, Koronyo *et al*. found high levels of unconjugated curcumin (400nM) without using β-glucuronidase [7]. This could be attributed to their different analysis methods: 1. wider/broader calibration settings. 2. internal standards without deuterated forms and validated in mouse. 3. acidification instead of freezing to stabilize samples.

Nevertheless, conjugated forms of curcumin are known to bind to amyloid as well following ex-vivo application to post-mortem brain sections [28]. And penetration of curcumin in the retina is suggested by our quantitative analysis where a general increase in fluorescence was observed after curcumin intake (data not shown), implying that curcumin with the capacity to bind fibrillar amyloid reached the retina. Despite 3-4 times higher plasma levels of curcumin conjugates using Theracurmin^®^ and Novasol^®^ compared to Longvida^®^ in this study, we found no difference in focal retinal hyperfluorescence between AD patients and controls.

The lack of between-group differences in our study could also be explained by absence of amyloid in the retina. The presence of (fibrillar) amyloid in the human retina is not unequivocally proven, as three labs showed positive staining with 6E10 and 12F4 antibodies interpreted as presence of retinal amyloid plaques in post-mortem retinas, while others were unable to replicate these findings [7, 10, 12, 13, 29, 30]. Methodological heterogeneity in post-mortem study methods might account for these discrepancies. Replication studies and harmonization of study methods are needed to overcome these discrepancies before the presence of retinal amyloid in AD can undoubtedly be confirmed. This is one of the key goals of ‘The Eye as a Biomarker for AD’ Personal Interest Area of the Alzheimer’s Association.

Three previous studies showed hyperfluorescence in AD cases after curcumin intake [7-9]. While these studies were of similar small sample size, AD biomarker confirmation was lacking, which is essential when relating retinal changes to AD, since other structures may cause changes in fluorescence as well. For example, retinal drusen, associated with macular degeneration, contain amyloid and other age-related deposits [31, 32]. This may account for the positive findings in Koronyo’s study where AD patients had a mean age of 76 years compared to 53 years in controls [7]. More work is needed to discriminate normal aging from pathological neurodegenerative changes in the retina underlying the observed changes in retinal fluorescence.

There are some limitations of this study, potentially explaining the negative findings. First, our scan timing might have been suboptimal for measuring curcumin bound to retinal amyloid resulting in impaired sensitivity. The optimal time point to measure retinal fluorescence is still to be determined. We based our scan timing on previously published pharmacokinetic curves of systemically available curcumin representing assumed peak levels of systemic curcumin, as has been shown to yield signal before [7, 22-25]. Second, the quantitative analysis could not be applied on the full cohort because of variation in imaging protocols. Also the fluorescent signal itself, is directly affected by illumination differences within images and between time points. These small alterations in scan quality may affect the co-registration of baseline and follow up scans and the signal itself. We applied several algorithms correcting for differences in illumination and scan quality. We are open to sharing our raw data, enabling application of other algorithms.

In conclusion, we found no differences in focal retinal hyperfluorescence between AD patients and controls pre- and post-curcumin, using Longvida^®^, a curcumin formulation previously used for this purpose. As we could not replicate previous findings with similar methods in our amyloid biomarker-confirmed cohort, we question whether focal retinal hyperfluorescence represents retinal amyloid, or rather age-related changes. Based on our analysis, retinal hyperfluorescence imaging using oral curcumin as labeling fluorophore is currently not ready for use as AD biomarker.

## Data Availability

Available upon reasonable request

## Authors’ contributions

| **Author** | **Contributions** |
| --- | --- |
| Jurre den Haan | Literature search, study design, data collection, data analysis, data interpretation, figures, writing-original draft. Verified data. |
| Frederique Hart de Ruyter | Data collection, data analysis, data interpretation, writing-review and editing |
| Benjamin Lochocki | Image analysis, quantitative data analysis, data interpretation, writing-review and editing |
| Maurice A.G.M. Kroon | Data collection plasma analysis, data analysis, data interpretation, writing-original draft |
| Marleen E. Kemper | Methodology plasma analysis, writing-review and editing, supervision |
| Charlotte E. Teunissen | Biomarker data interpretation, writing-review and editing |
| Bart van Berckel | Biomarker data interpretation, writing-review and editing |
| Philip Scheltens | Funding acquisition, writing-review and editing |
| Jeroen J. Hoozemans | Data interpretation, writing-review and editing |
| Aleid Kreeke | Data collection |
| Frank D. Verbraak | Methodology, study design, image data collection, data analysis, data interpretation, writing-review&editing. Verified data. |
| Johannes F. de Boer | Funding acquisition, methodology, study design, data interpretation, writing-review and editing. Verified data. |
| Femke H. Bouwman | Funding acquisition, study design, data analysis, data interpretation, writing-original draft. Verified data. |

## cknowledgements/Funding

We gratefully acknowledge financial support from Stichting Alzheimercentrum VUMC, Alzheimer Nederland, the Dutch Technology Foundation STW (grant number 13935), part of the Netherlands Organization for Scientific Research (NWO), and which is partly funded by the Ministry of Economic Affairs, ISAO (grant number 14518).

## Data availability

Raw imaging and clinical data is available upon request

## Supplemental material

**Supplemental table 1.** Demographics for each individual participant.

| # | Dx | Age | Sex | MMSE | CSF analysis |  |  |  | CSF assay | Amyloid-PET | Curcumin |
| --- | --- | --- | --- | --- | --- | --- | --- | --- | --- | --- | --- |
| | | | | | Aβ | Tau | pTau | pTau/Aβ | | | |
| 1 | AD | 64 | M | 20 | 778 | 551 | 60 | 0.077 | Elecsys | n.a. | Longvida |
| 2 | AD | 57 | M | 23 | 342 | 108 | 10 | 0.029 | Elecsys | n.a. | Longvida |
| 3 | AD | 70 | F | 30 | 511 | 211 | 21 | 0.041 | Elecsys | n.a. | Longvida |
| 4 | AD | 72 | M | 22 | 805 | 316 | 35 | 0.043 | Elecsys | n.a. | Longvida |
| 5 | CN | 54 | F | 30 | 641 | 208 | 19 | 0.030 | Elecsys | (-) 18F-Florbetapir | Longvida |
| 6 | AD | 69 | M | 24 | 322 | 340 | 34 | 0.106 | Elecsys | n.a. | Longvida |
| 7 | AD | 56 | F | 21 | 746 | 215 | 23 | 0.031 | Elecsys | n.a. | Longvida |
| 8 | CN | 57 | M | 28 | 1323 | 136 | 11 | 0.008 | Elecsys | (-) 18F-Florbetapir | Longvida |
| 9 | AD | 59 | F | 19 | 337 | 156 | 17 | 0.050 | Elecsys | n.a. | Longvida |
| 10 | CN | 89 | F | 28 | n.a. | n.a. | n.a. | n.a. | n.a. | (-) 18F-Flutemetamol | Longvida |
| 11 | CN | 70 | F | 29 | n.a. | n.a. | n.a. | n.a. | n.a. | (-) 18F-Flutemetamol | Longvida |
| 12 | AD | 75 | M | 28 | n.a. | n.a. | n.a. | n.a. | n.a. | (+) 18F-Florbetaben | Longvida |
| 13 | AD | 79 | M | 27 | n.a. | n.a. | n.a. | n.a. | n.a. | (+) 18F-Florbetaben | Longvida |
| 14 | AD | 81 | F | 26 | n.a. | n.a. | n.a. | n.a. | n.a. | (+) 18F-Florbetaben | Longvida |
| 15 | CN | 80 | M | 29 | n.a. | n.a. | n.a. | n.a. | n.a. | (-) 18F-Florbetapir | Longvida |
| 16 | AD | 77 | M | 25 | n.a. | n.a. | n.a. | n.a. | n.a. | (+) 18F-Flutemetamol | Longvida |
| 17 | AD | 78 | M | 27 | n.a. | n.a. | n.a. | n.a. | n.a. | (+) 18F-Florbetaben | Longvida |
| 18 | AD | 81 | M | 27 | n.a. | n.a. | n.a. | n.a. | n.a. | (+) 18F-Florbetaben | Longvida |
| 19 | CN | 66 | F | 29 | n.a. | n.a. | n.a. | n.a. | n.a. | (-) 18F-Flutemetamol | Longvida |
| 20 | CN | 82 | F | 29 | n.a. | n.a. | n.a. | n.a. | n.a. | (-) 18F-Flutemetamol | Longvida |
| 21 | CN | 79 | M | 30 | 1328 | 279 | 26 | 0.020 | Elecsys | (-) 18F-Florbetapir | Longvida |
| 22 | CN | 78 | F | 27 | n.a. | n.a. | n.a. | n.a. | n.a. | (-) 18F-Flutemetamol | Longvida |
| 23 | CN | 84 | M | 29 | 1498 | 364 | 33 | 0.022 | Elecsys | (-) 18F-Florbetapir | Longvida |
| 24 | AD | 77 | F | 23 | 642 | 460 | 45 | 0.070 | Elecsys | (+) 18F-Flutemetamol | Longvida |
| 25 | CN | 66 | F | 30 | n.a. | n.a. | n.a. | n.a. | n.a. | (-) 18F-Flutemetamol | Longvida |
| 26 | CN | 76 | F | 29 | n.a. | n.a. | n.a. | n.a. | n.a. | (-) 18F-Flutemetamol | Longvida |
| 27 | AD | 67 | M | 21 | 548 | 394 | 35 | 0.064 | Innotest | (+) 18F-Florbetapir | Theracurmin |
| 28 | AD | 61 | M | 20 | 738 | 307 | 31 | 0.042 | Innotest | (+) 18F-Florbetaben | Theracurmin |
| 29 | CN | 63 | F | 30 | n.a. | n.a. | n.a. | n.a. | n.a. | (-) 18F-Florbetapir | Theracurmin |
| 30 | AD | 64 | M | 18 | 291 | 847 | 67 | 0.231 | Innotest | (+) 18F-Florbetaben | Theracurmin |
| 31 | AD | 66 | M | 28 | 637 | 282 | 24 | 0.037 | Innotest | (+) 18F-Florbetaben | Theracurmin |
| 32 | CN | 51 | F | 30 | 1604 | 110 | 7 | 0.004 | Innotest | n.a. | Theracurmin |
| 33 | AD | 57 | F | 19 | 843 | 587 | 48 | 0.057 | Innotest | (+) 18F-Florbetaben | Theracurmin |
| 34 | AD | 61 | M | 25 | 456 | 367 | 40 | 0.088 | Innotest | (+) 18F-Florbetaben | Theracurmin |
| 35 | AD | 54 | M | 24 | 521 | 322 | 30 | 0.058 | Innotest | (+) 18F-Florbetaben | Theracurmin |
| 36 | AD | 69 | M | 25 | 693 | 318 | 35 | 0.051 | Innotest | (+) 18F-Florbetaben | Novasol |
| 37 | AD | 65 | F | 24 | 534 | 477 | 46 | 0.086 | Innotest | (+) 18F-Florbetaben | Novasol |
| 38 | AD | 60 | M | 18 | 497 | 232 | 23 | 0.047 | Innotest | n.a. | Novasol |
| 39 | AD | 56 | F | 23 | 892 | 486 | 33 | 0.037 | Innotest | n.a. | Novasol |
| 40 | AD | 55 | F | 24 | 525 | 270 | 25 | 0.048 | Innotest | n.a. | Novasol |

Cohort characteristics of participants enrolled in cohort 1 (Longvida®), cohort 2
(Theracurmin®) and cohort 3 (Novasol®). CSF data is presented as pg/ml. pTau/Aβ
ratio $\geq 0.020$ was considered as AD profile.<sup>17</sup> Abbreviations: # = Participant Number,
Dx =Diagnosis, MMSE = Mini-Mental State Examination, CSF = Cerebrospinal Fluid,
Aβ = Amyloid-beta, pTau = Phosphorylated Tau, + = positive, - = negative, n.a. = not
available.

**Supplemental Figure 1.**
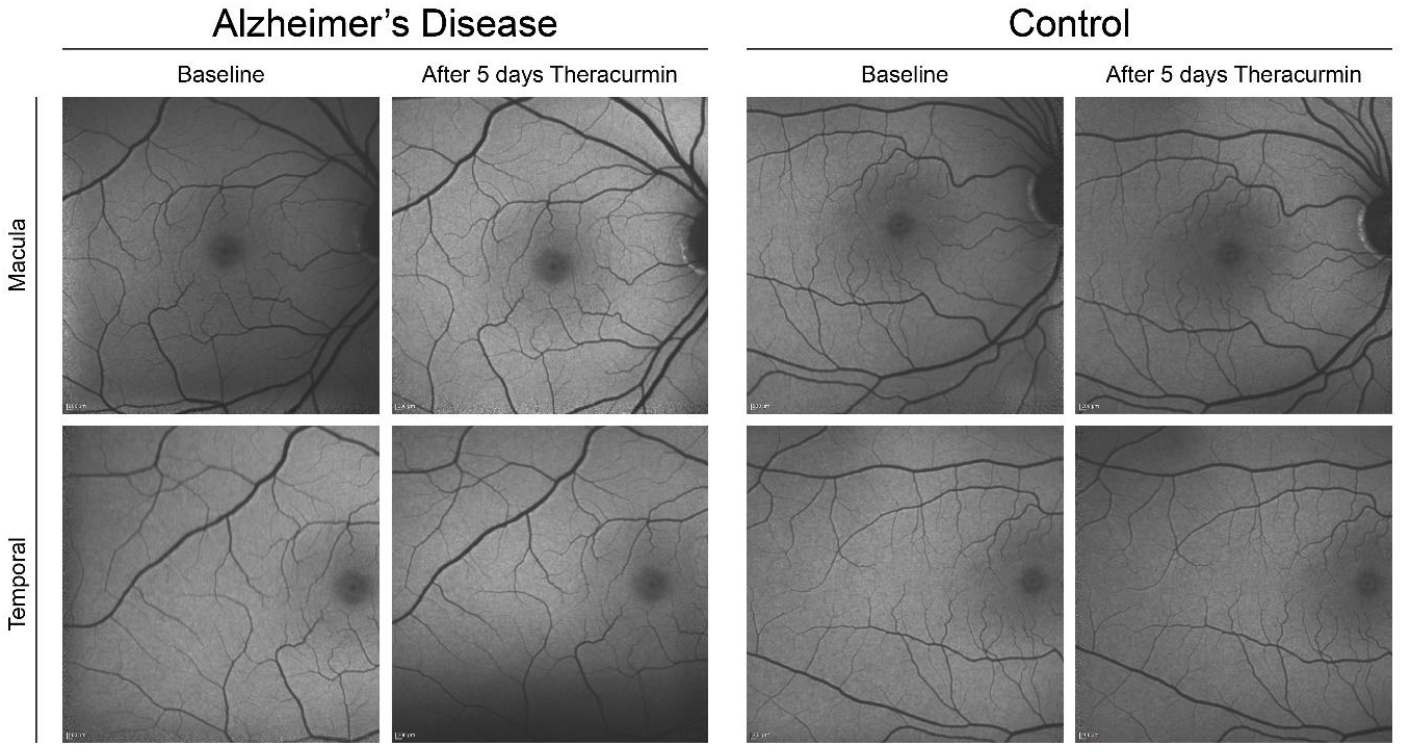
Pre- and post-curcumin retinal fluorescence images for AD patients and controls using Novasol^®^. Pre-and post-curcumin retinal fluorescence images with a 55-degree lens using blue auto fluorescence (λ=486nm) in 6 retinal regions in a representative Alzheimer’s disease (AD) patient. Abbreviations: TS= temporal-superior, S= superior, T= temporal, M= macula, TI= temporal-inferior, I= inferior.

**Supplemental Figure 2.**
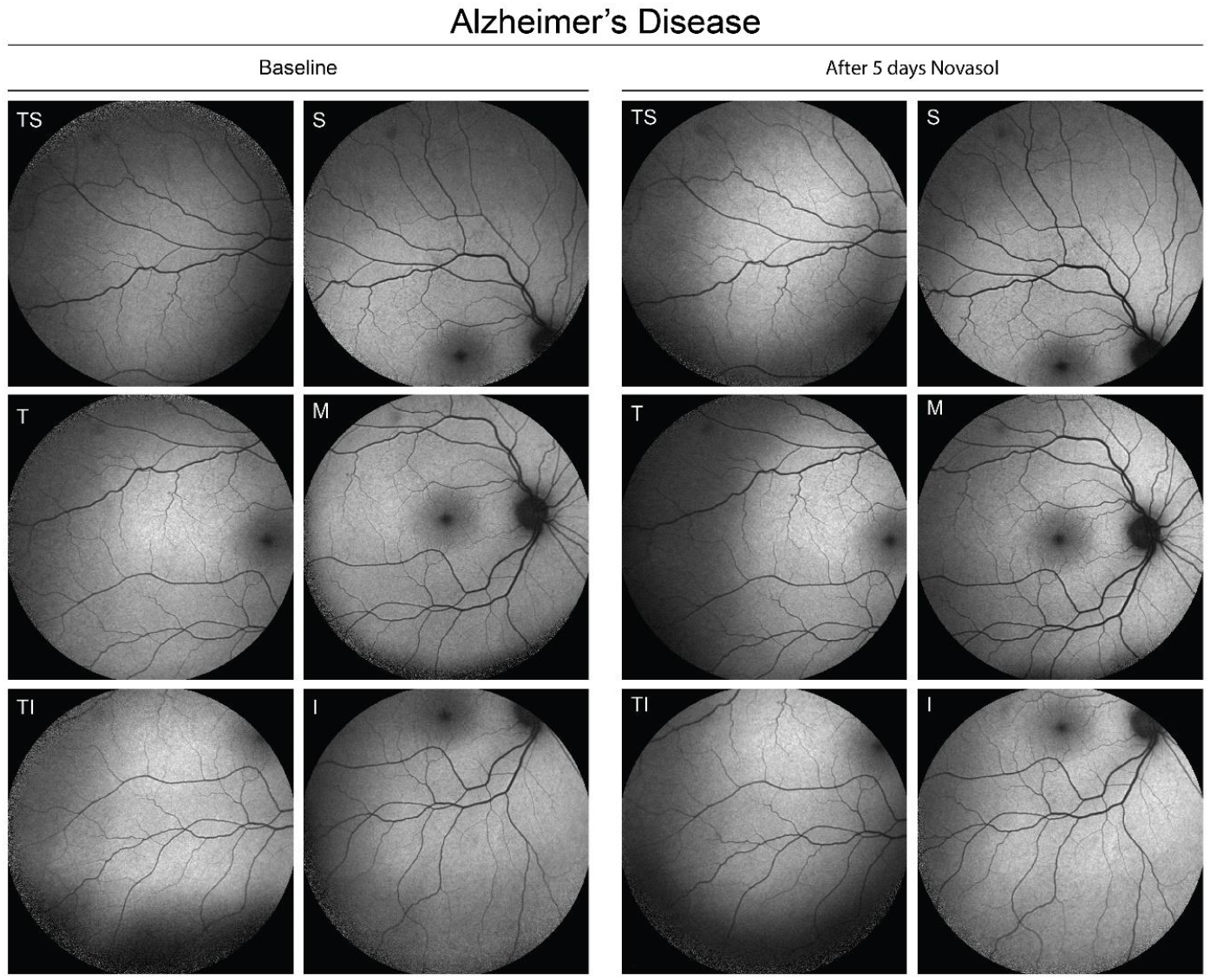
Pre- and post-curcumin retinal fluorescence images for AD patients and controls using Theracurmin^®^. Pre-and post-curcumin retinal fluorescence images with a 30-degree lens using blue auto fluorescence (λ=486nm) in 2 retinal regions in a representative Alzheimer’s disease (AD) patient and control. Abbreviations: M= macula, T= temporal.

